# Development and Validation of OncoOrigin: An Integrative AI Tool for Primary Cancer Site Prediction

**DOI:** 10.1101/2024.11.29.24318189

**Authors:** Petar Brlek, Luka Bulić, Nidhi Shah, Parth Shah, Dragan Primorac

**Author notes:** These authors contributed equally to this work (Petar Brlek, Luka Bulić). Corresponding author: Petar Brlek St. Catherine Specialty Hospital, Branimirova street 71E, 10000 Zagreb, Croatia.

## Abstract

**Importance:** Cancers of unknown primary origin (CUPs) represent a significant diagnostic and therapeutic challenge in the field of oncology. With the limitations of current diagnostic tools in these cases, novel approaches must be brought forward to improve treatment outcomes for these patients.

**Objective:** The objective of this study was to develop a machine-learning-based software for primary cancer site identification (OncoOrigin), based on genetic data acquired from tumor DNA sequencing.

**Design:** By design, this was an *in silico* diagnostic study.

**Setting:** This study was conducted using data from the cBioPortal database (accessed on 21 September 2024) and several data processing and machine-learning Python libraries.

**Participants:** This study involved over 20,000 tumor samples with information on patient age, sex, and the presence of genetic variants in over 600 genes.

**Main Outcomes and Measures:** The main outcome of interest in this study was machine-learning-based discrimination between cancer type classes, based on the provided data. Model quality was assessed by train set cross-validation and evaluation on a segregated test set. Finally, the optimal model was incorporated with a graphical user interface into the OncoOrigin software. Feature importances for class discrimination were also determined on the optimal model.

**Results:** Out of the four tested machine-learning estimators, the XGBoostClassifier-based model proved superior on test set evaluation, with a top-2 accuracy of 0.91 and ROC-AUC of 0.97. Class sensitivity values for prostate cancer, breast cancer, melanoma, and colorectal cancer were over 0.85, while all class specificity values were equal to or higher than 0.95. The top 3 significant features were patient sex, and genetic alterations in the *APC* and *KRAS* genes.

**Conclusions and Relevance:** In this study, we have successfully developed a machine-learning-based software for primary cancer site identification with high-quality evaluation metrics. Through simple clinical implementation, such a tool has the potential to significantly improve the diagnostics and treatment outcomes for patients suffering from CUP.

**KEY POINTS:** 

**Question:** Can a machine-learning-based tool (OncoOrigin) accurately predict the primary cancer site from tumor DNA sequencing data in patients with cancers of unknown primary origin (CUP)?

**Findings:** In this *in silico* study, the XGBoostClassifier model demonstrated high predictive performance, achieving a top-2 accuracy of 0.91 and ROC-AUC of 0.97 for the classification of ten cancer types using genetic data from over 20,000 tumor samples.

**Meaning:** The OncoOrigin software shows potential for improving the diagnosis and treatment of CUP by providing accurate primary cancer site identification based on genetic data.

## INTRODUCTION

Cancers of unknown primary origin (CUPs) make up 5% of all malignant conditions worldwide. They are characterized by their aggressiveness, fast progression, and poor prognosis with alarmingly high mortality rates. Up to 84% of patients have fatal outcomes within the first year after diagnosis and represent some of the most diagnostically and therapeutically challenging cases in the field of oncology. A lack of information regarding the primary origin limits treatment possibilities as oncological protocols are largely based on the primary tumor site. Traditional diagnostic methods, such as immunohistochemistry using specific markers, have limited diagnostic power, especially in poorly differentiated tumors, and are often not sufficient for primary site verification [1, 2].

Tumor molecular profiling with novel next-generation sequencing (NGS) methods, such as whole genome sequencing (WGS), has opened up the possibility of detecting all potential germline and somatic genetic alterations in tumor tissue. This enables more precise molecular classification of tumors and their personalized treatment [3]. Scientific efforts have been made to use genetic profiling for precise molecular classification of tumors. One such example is using single-cell transcriptomics and spatial proteomics with the purpose of improving treatment outcomes [4].

In recent years, machine learning has become a robust tool in biomedical research, due to its ability to efficiently process large quantities of data [5]. Certain studies have endeavored to combine genetic testing and machine learning with the purpose of primary site identification [6]. While promising results have been demonstrated, further research and validation are required prior to the implementation of such technology into clinical practice.

The aim of our study was to train a primary cancer site predictor by combining genetic data from over 20,000 tumor samples and different machine-learning algorithms. Following evaluation, the optimal model would be fitted into a simple software application which would allow for easy practical implementation. Such a tool could offer a new and robust diagnostic option for CUP cases and potentially significantly improve treatment outcomes for these patients, and in the future, enable the detection of the primary tumor site based on the analysis of liquid biopsies without the need for invasive and harmful medical procedures.

## METHODOLOGY

### Study design

By design, this was an *in silico* study that involved the curation, processing, and utilization of publicly available data to develop a new machine-learning model (Figure 1). All of the data and code used in this study are publicly available (stated in the Data sharing statement) and the study design was approved by an institutional ethics committee (stated in the Ethical statement).

**Figure 1.**
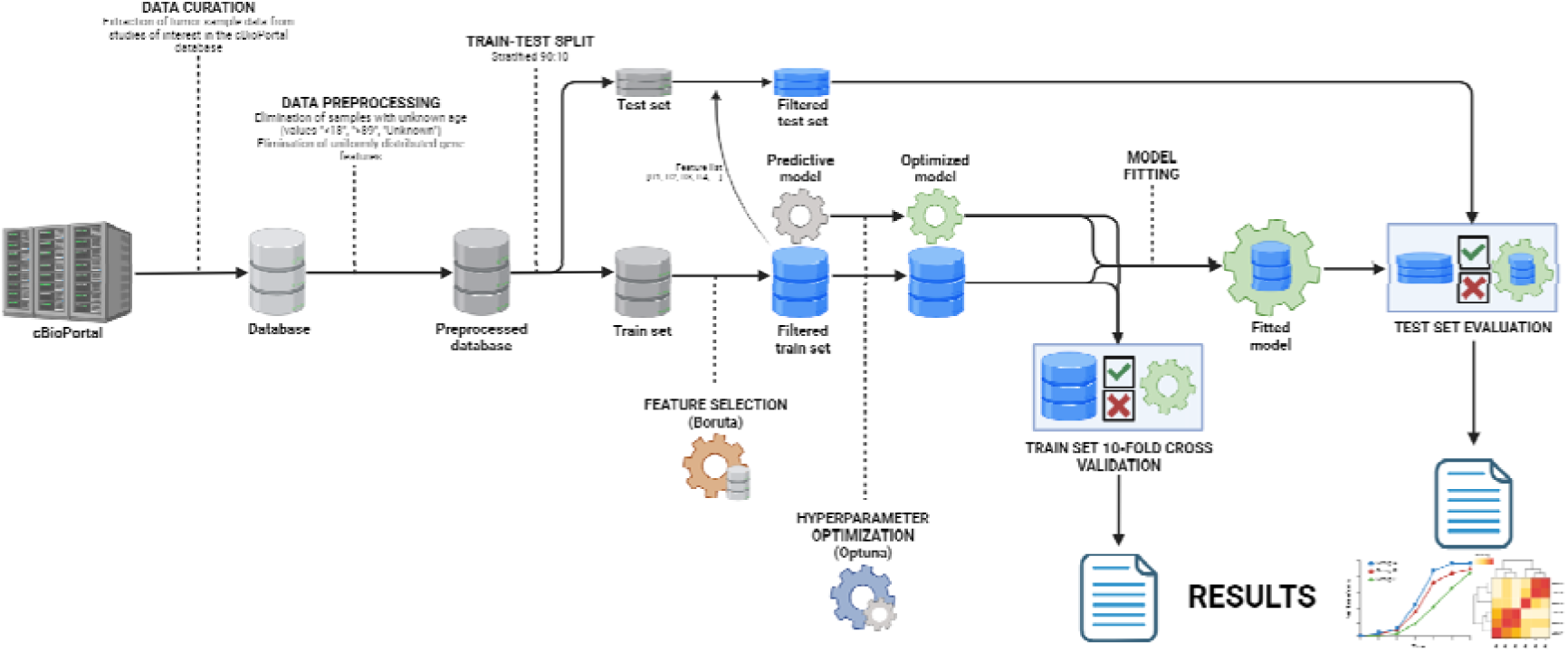
Data curation and model development flow chart (created with Biorender.com).

### Data curation

The data were extracted from cBioPortal (https://www.cbioportal.org/, accessed on 21 September 2024), a comprehensive database for cancer genomics [7, 8, 9]. cBioPortal contains clinical and genomic data from the AACR Project GENIE® (Genomic Evidence Neoplasia Information Exchange). AACR Project GENIE (https://genie.cbioportal.org/, accessed on 21 September 2024) is a publicly accessible registry of real-world genomic data created through collaboration between 19 leading international cancer centers [10]. With approximately 287,000 sequenced samples from over 191,000 patients, it is one of the largest publicly available cancer genomic datasets, offering a vast resource for understanding the molecular underpinnings of various cancers. From this extensive dataset, 21,016 samples were selected through a rigorous filtering process for use in machine learning model development. The filtering process involved key criteria to refine the dataset. Only one sample per patient was included to avoid multiple sequencing events for the same metastatic tumor. The selection included 62.3% female (13,085 samples) and 37.7% male (7,931 samples) patients. The patient’s age at the time of sequencing was also utilized from the dataset. The virtual study created included 19 different subtypes of metastatic tumors.

Data for 665 genes were extracted for each sample, selected according to previously published validated panels as part of the implementation of whole-exome sequencing for tumor tissue analysis [11]. For the processing and utilization of data on single nucleotide variants (SNVs), copy number variants (CNVs), and structural variants to develop machine learning models, a binary system was employed. The presence of an SNV, CNV, or structural gene alteration was coded as 1, while the absence of these changes was coded as 0.

### Data preprocessing

Initial preprocessing included elimination of samples with unknown patient age (classified as “Unknown”, “<18”, or “>89”) and samples with Not-a-Number (NaN) values. Furthermore, elimination of gene features with no detected variants across all samples was done. Additionally, the “Hepatobiliary Cancer” and “Pancreatic Cancer” classes were merged into the “Hepatobiliary/Pancreatic Cancer” class and the “Small Cell Lung Cancer” and “Non-Small Cell Lung Cancer” classes were merged into the “Lung Cancer” class. The output vector consisted of ten classes, “Colorectal Cancer”, “Ovarian Cancer”, “Melanoma”, “Breast Cancer”, “Lung Cancer”, “Thyroid Cancer”, “Prostate Cancer”, “Hepatobiliary/Pancreatic Cancer”, “Bladder Cancer”, and “Endometrial Cancer”.

### Model development

Four robust ensemble/gradient boosting machine-learning algorithms, based on decision trees, were used for the prediction of cancer type – RandomForestClassifier (RFC), XGBoostClassifier (XGBC), CatBoostClassifier (CBC), and ExtraTreesClassifier (ETC). The preprocessed database was initially split into train and test sets in a 90:10 size ratio, using stratification with respect to the predicted class. Feature selection among the gene features was done on the train set, using the Boruta algorithm with an optimized XGBC estimator. Secondly, hyperparameter optimization based on the train set was done for each of the four algorithms, using the Optuna library. Finally, the models were initialized with optimal hyperparameters and fitted with the filtered train set. After the evaluation stage, the best estimator was chosen and fitted with the entire preprocessed database. This model was used for the development of the OncoOrigin software and feature importance analysis.

### Model evaluation

After fitting, each of the estimators was evaluated based on several metrics. All metrics were determined using the one-versus-rest setting for multiclass classification. The first step of evaluation was stratified 10-fold cross-validation on the train set, after feature selection and hyperparameter optimization. In this step, mean accuracy scores, mean weighted F1 scores, and mean weighted receiver operating characteristic area under curve (ROC-AUC) scores were determined from each of the ten folds. Additionally, 95% confidence intervals were determined for these metrics. The second step involved performance evaluation on the segregated test set. For this analysis, accuracy scores, top-2 accuracy scores, weighted F1 scores, weighted ROC-AUC scores, weighted precision-recall area under curve (PR-AUC) scores, and class-specific sensitivity and specificity values were determined. Top-2 accuracy represents accuracy but the prediction is considered correct if the true value is one of the two most probable predicted classes. Additionally, confusion matrices and class-specific PR and ROC curves were determined for each estimator based on test set evaluation.

### User interface development

The final fitted XGBC model was exported and packaged into a software with a graphical user interface (GUI) - OncoOrigin (Supplemental figure S1). In the software, the user input consists of the patient’s age at time of sequencing, the patient’s sex, and genes in which genetic variants were discovered after tumor sequencing. The interface outputs the predicted class, as well as prediction confidence and probabilities for all other classes relative to the predicted class. The prediction confidence is determined by the relative probability *p_2_* of the second most probable class (*p_2_* ≥ 0.8 for very low, 0.8 > *p_2_*≥ 0.6 for low, 0.6 > *p_2_* ≥ 0.4 for moderate, 0.4 > *p_2_*≥ 0.2 for high, and *p_2_* < 0.2 for very high).

### Programming implementation

All steps involving machine learning model training and evaluation, as well as GUI development, were done in the Python/Jupyter Notebook programming language (v3.12.3) [12]. Several Python libraries were used, including Pandas (v2.2.2), NumPy (v1.26.0), MatPlotLib (v3.8.0), Seaborn (v0.13.2), SciPy (v1.14.0), Scikit-Learn (v1.5.1), XGBoost (v2.1.0), CatBoost (v1.2.5), Boruta (v0.3), and Optuna (v3.6.1) [13–22]. Development of the OncoOrigin GUI was done using the Tkinter (v8.6.13) library [23]. Random state variables throughout the code were defined by a chosen value. Still, due to inherent non-determinism in some of the stated libraries, different runs of the code still did not produce the exact same results. However, noticed deviations in evaluation metric values due to non-determinism were negligible.

## RESULTS

### Data preprocessing

After data extraction from the cBioPortal database had been completed, data on sex, age at time of sequencing, and genetic variant presence for 665 genes across 21,016 patient tumor samples were selected (Supplemental table S2). Elimination of uniformly distributed genes produced the database for the modeling pipeline, containing the features of sex, age, and genetic variant presence for 566 genes and 20710 samples. After feature selection, 345 genetic features remained in the database, along with the sex and age features (Supplemental table S3). Class frequencies in the output vector were 0.14, 0.09, 0.08, 0.22, 0.22, 0.02, 0.07, 0.11, 0.02, and 0.02 for colorectal cancer, ovarian cancer, melanoma, breast cancer, lung cancer, thyroid cancer, prostate cancer, hepatobiliary/pancreatic cancer, bladder cancer, and endometrial cancer respectively.

### Model evaluation

After feature selection, hyperparameter optimization, and model fitting were complete, evaluation metrics were determined for each of the four estimators. This included metrics received from train set cross-validation and evaluation based on the test set (Table 1). The results demonstrated the XGBC model as the superior model in both train set cross-validation and test set evaluation. The CBC model showed only slightly poorer performance metrics, while the ETC and RFC showed significantly poorer performance metrics. Interestingly, the ETC model, while poorer in overall quality, showed the highest sensitivity values for ovarian cancer, thyroid cancer, bladder cancer, and endometrial lower cancer.

**Table 1.**
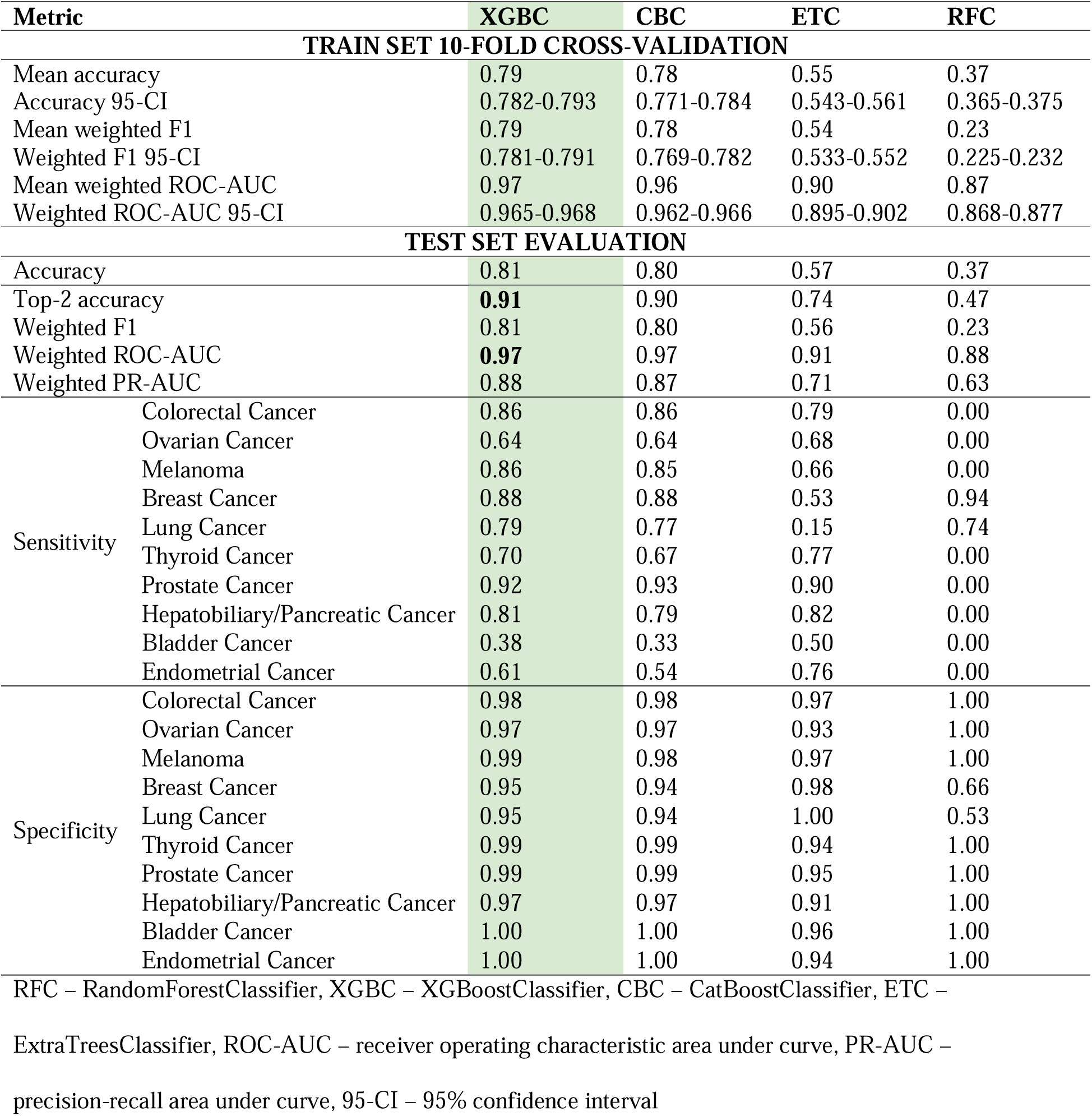
Evaluation metrics for each estimator.

### Class-specific evaluation and feature analysis

Confusion matrices were determined for each of the models to better evaluate how each class impacted the overall predictive power (Figure 2). The RFC confusion matrix showed that the model predicted almost all samples as either breast cancer or lung cancer. The ETC confusion matrix showed a somewhat better result, with a higher ability to detect less represented classes, but a poor ability to detect the lung cancer class. Finally, the XGBC and CBC confusion matrices had a very similar layout. Hotspots were noticed in both fields pertaining to breast cancer and ovarian cancer, indicating a difficulty in differentiating between the two. ROC curves were in line with the model metrics (Figure 3). In terms of class, the bladder cancer and lung cancer ROC curves visibly stood out with lower AUCs in all models, while the prostate cancer ROC curves were consistently superior across all models. A similar trend was noted in the precision-recall curves (Supplemental figure S4). The feature importance analysis revealed the top 20 features for primary site prediction (Figure 4). The greatest importance score was calculated for the patient sex feature, followed by *APC*, and *KRAS*.

**Figure 2.**
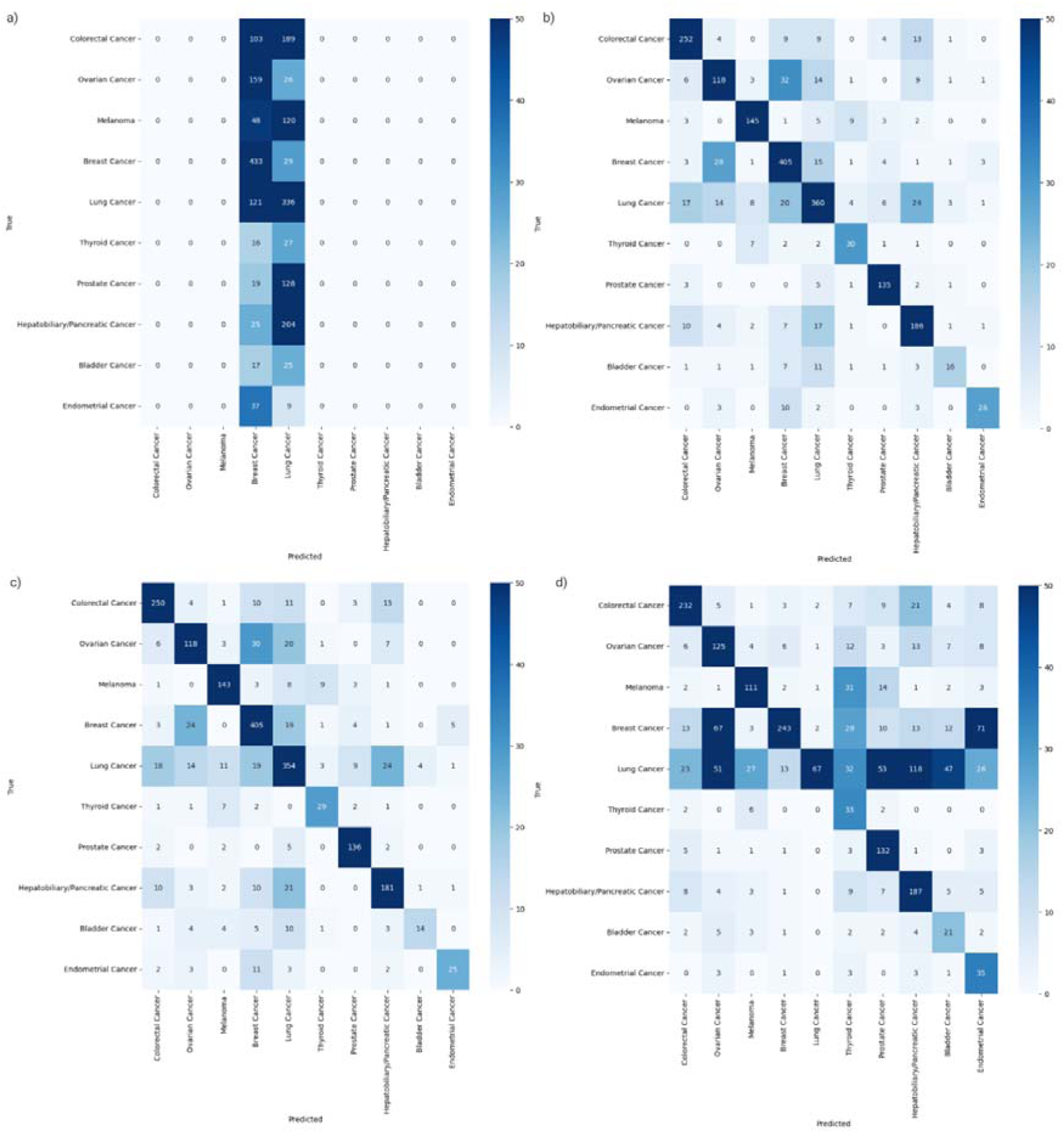
Confusion matrices of the RFC model (a), XGBC model (b), CBC model (c), and ETC model (d).

**Figure 3.**
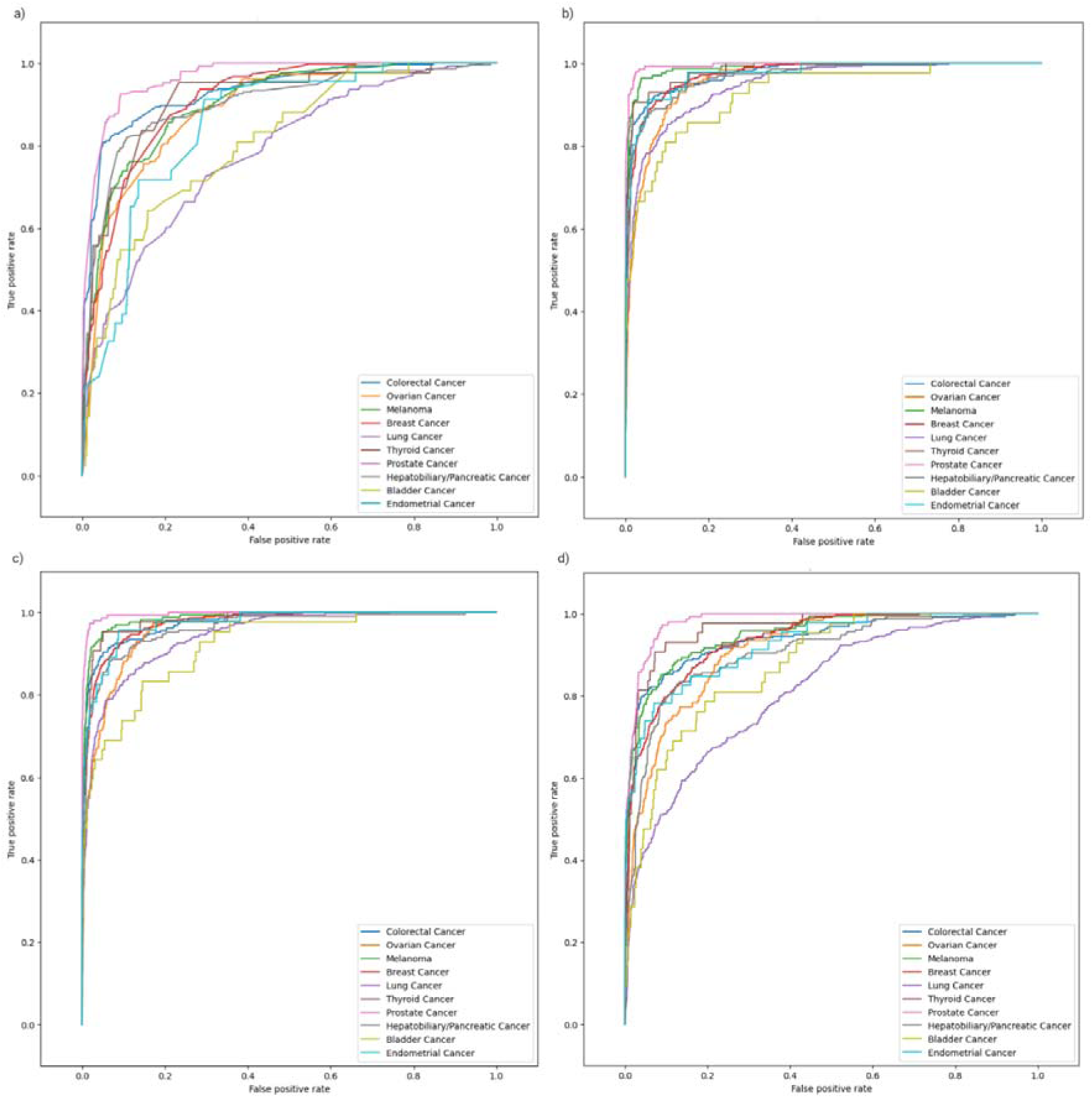
Class-specific ROC analysis curves for the RFC model (a), XGBC model (b), CBC model (c), and ETC model (d).

**Figure 4.**
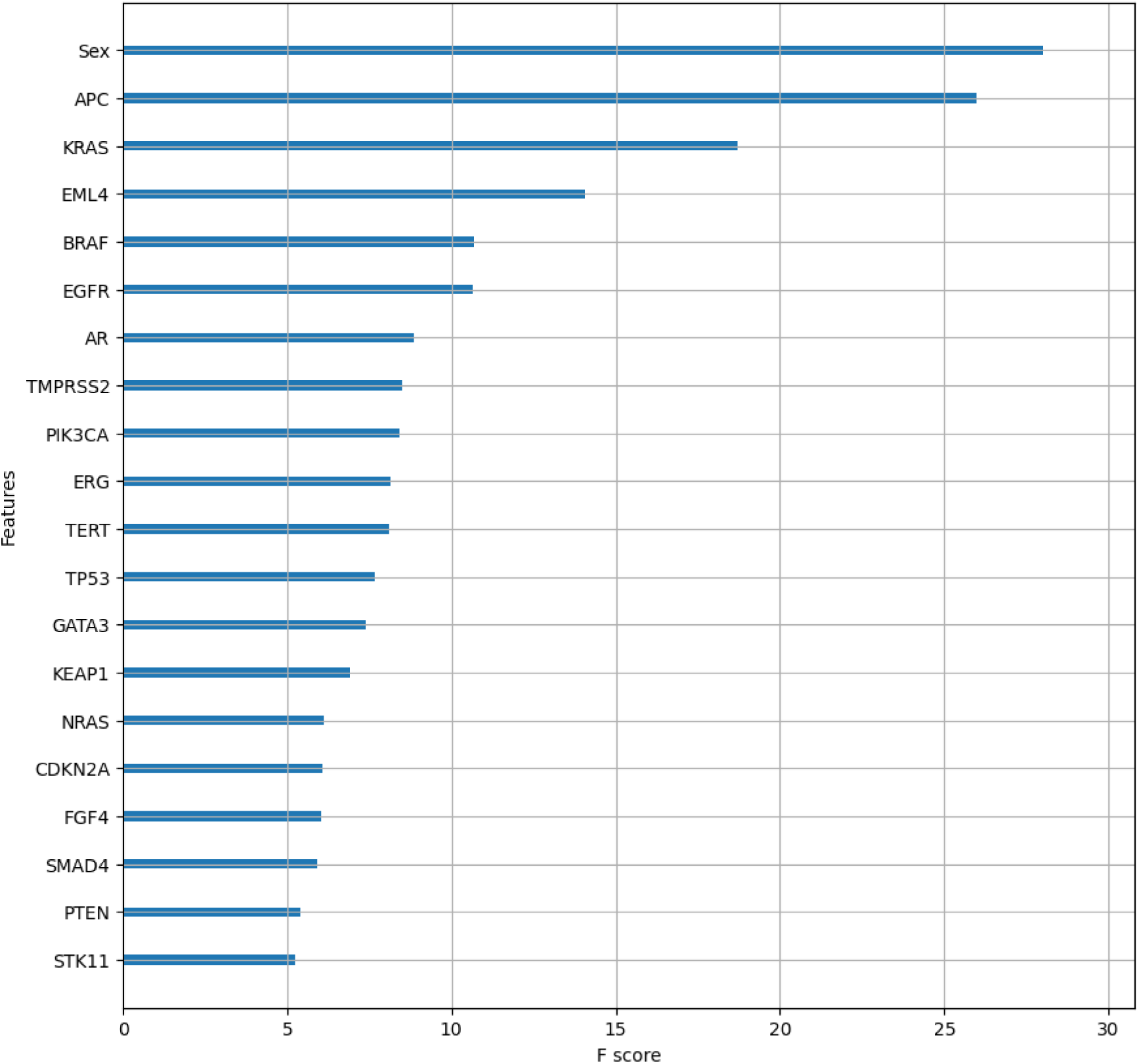
Feature importance analysis of the top 20 features based on the optimal XGBC model.

## DISCUSSION

### Software quality evaluation and comparison

After evaluation and interpretation of the produced results, the XGBoostClassifier was identified as the optimal estimator for OncoOrigin, with an accuracy of 0.81, top-2 accuracy of 0.91, F1-score of 0.81, PR-AUC score of 0.88, and ROC-AUC score of 0.97. As the software provides the user with not only the prediction but also insight into the likelihood of the other classes, clinicians will always be informed whether a second class is highly likely. As this can be clinically actionable information, we deemed the top-2 accuracy metric relevant for our model. When observing specific classes, sensitivity values ≥0.79 were found for breast cancer, lung cancer, colorectal cancer, prostate cancer, melanoma, and hepatobiliary/pancreatic cancer, while the other classes had lower sensitivity values. This trend generally corresponds to the quantitative representation of each class in the total population sample, determined by frequency analysis. Where class specificity values are concerned, all values were ≥0.95.

The performance metrics of our model can be compared to other relevant literature for this context. In the paper published by Moon I et al., the authors also proposed a machine-learning model for primary cancer site detection [6]. While the source of data, number of samples used for training and testing, and feature definition and representation were different, the authors reported a weighted F1-score of 0.784 for classification based on 22 cancer types and an F1-score of 0.806 for classification based on 13 cancer types. Our OncoOrigin model measures up to these metrics with a weighted F1-score of 0.81 for classification based on 10 cancer types. Furthermore, while the authors used different feature selection and hyperparameter optimization methods, they too used the XGBoostClassifier algorithm as the classification estimator. In an article by Bicakci N, the effectiveness of the F-18 FDG PET-CT scan in CUP cases was evaluated [24]. The primary tumor was correctly determined in 41% of patients. The rest of the patients either did not have matching PET-CT or histopathology findings, or failed to have their primary tumor differentiated by both methods. Another study, published by Lu MY et al., constructed a deep-learning model to identify primary cancer sites based on histopathological image processing [25]. In their article, the authors reported their model achieved an accuracy of 0.834 when evaluated on the segregated test set. Once again, this is comparable with the 0.81 accuracy which OncoOrigin achieved on our segregated test set.

When comparing our software with the literature, the main strength of OncoOrigin is its integration with a graphical user interface. Normally, interactions with developed machine-learning models require a background in AI programming and data science. However, for clinicians and other medical experts, this might be an impractical skillset to acquire and maintain [26]. For this reason, a simple user interface which communicates data between the model and the user greatly facilitates its potential for implementation in clinical practice.

### Software limitations

While quality results have been achieved in the evaluation of OncoOrigin, the model does have several limitations. Firstly, the model has a total of 347 input features, the values of which should be known upon using the software to receive a prediction with an accuracy level corresponding to the one presented in this article. This entails the sequencing of 345 genes, which may be technically difficult, depending on the available technology for any given user. Secondly, due to the format of the extracted data, the model has been trained with age values ranging from 18 to 89. For this reason, the age input in the software has been limited to this range. Thirdly, while the model did use data combined from multiple studies, this data was still extracted from a single source which leaves potential for a certain level of data bias and risk of overfitting. Another limitation regarding the data itself is underrepresentation of certain cancer types, which likely negatively impacted the ability of the model to identify them. Finally, the model is limited to predicting primary sites among the ten cancer type classes included in the study.

### Practical applications of OncoOrigin in precision oncology

The possibility of tumor sequencing and primary cancer site prediction in CUP cases holds the potential for significant treatment benefits in the context of targeted oncological therapy. Precision medicine has revolutionized oncological therapy by enabling the implementation of targeted treatments based on specific molecular biomarkers of tumors [27]. Modern molecular biology techniques, such as whole genome sequencing (WGS) and whole exome sequencing (WES) of tumor tissue, facilitate the determination of the molecular profile of tumors and the application of therapies that act on specific molecular targets within the tumor tissue [3]. A key tool in this approach is tumor molecular profiling, which involves the genetic analysis of tumor tissue embedded in paraffin and/or circulating tumor DNA (ctDNA) obtained from liquid biopsy samples. The analysis of ctDNA from a patient’s blood represents a significant advancement in oncology, especially in cases where traditional tissue biopsies are not feasible [28]. These advancements in oncological diagnostics form the basis for the introduction of molecular classification of neoplasms aimed at targeted treatment of secondary tumors of unknown primary origin. The further development of the OncoOrigin platform, in addition to the precise detection of the primary tumor site, opens up extensive possibilities for the molecular classification of secondary neoplasms based on results obtained through NGS methods. This advancement will pave the way for the diagnosis and precise treatment of CUP. Our planned further research will validate the *in vivo* data obtained from the genetic analysis of tumor tissue embedded in paraffin and ctDNA derived from liquid biopsy samples. Should the quality of OncoOrigin the model hold its standard when tested on data obtained from liquid biopsy, we open the possibility for early detection and treatment of metastatic cancers without relying on standard invasive methods such as surgery and PET-CT scans, which expose patients to additional ionizing radiation that can damage healthy cells [29]. Furthermore, in the case that more than one primary cancer site is likely, our software offers this information to the clinician. As multiple primary cancers can be managed with a combined regimen, this can in certain cases be actionable information [30].

### Conclusion and future directions

The results presented in this study present OncoOrigin as a robust tool for primary cancer site identification, with great potential application in CUP cases. While the software has certain limitations, it achieved excellent scores on the most frequent cancer types with significant metastatic potential. Furthermore, the software does not only offer the prediction of the model itself but information on the probability of each tumor type included. This provides clinicians with a greater amount of relevant and actionable information than models that offer the predicted class alone. As all the training and evaluation in this study were done *in silico*, the next phase of OncoOrigin validation will be multi-centric evaluation on patients from our population diagnosed with metastatic cancer with a known primary site. Finally, provided we achieve an equal standard of results, the third phase will involve prediction of the primary cancer site in CUP cases and an analysis of outcome benefit. Our end goal with OncoOrigin is its clinical implementation in precision oncology and a significant change in the clinical paradigm of CUP.

## Supporting information

Supplemental figure S1. OncoOrigin graphical user interface

Supplemental table S2. List of all genes included in data extraction from cBioPortal

Supplemental table S3. List of all genes included in modeling after preprocessing and feature selection

Supplemental figure S4. Class-specific precision-recall analysis curves for the RFC model (a), XGBC model (b), CBC model (c), and ETC model (d)

## Data Availability

The data used in this study is publicly available as part of a virtual study in the cBioPortal system, available at the following link: https://genie.cbioportal.org/study?id=670d2529854f636a3863065d.
The source code files used for data management, model training and evaluation, and user interface development are publicly available as part of an active GitHub repository, available at the following link: https://github.com/lbulic1003/OncoOrigin (Source code/v1.0.0).

https://github.com/lbulic1003/OncoOrigin

## Acknowledgements

We thank the International Society for Applied Biological Sciences (ISABS) for their continued support.

## Data sharing statements

The data used in this study is publicly available as part of a virtual study in the cBioPortal system, available at the following link: https://genie.cbioportal.org/study?id=670d2529854f636a3863065d.

The source code files used for data management, model training and evaluation, and user interface development are publicly available as part of an active GitHub repository, available at the following link: https://github.com/lbulic1003/OncoOrigin (Source code/v1.0.0).

## Ethical statement

This study was approved by the St. Catherine Specialty Hospital ethics committee (Sep 2024, 24/8-I) and was conducted in accordance with the Helsinki declaration.

## Reporting guidelines

This manuscript was written in accordance with the TRIPOD+AI checklist.

## Study registration and protocol

No registration was done for this study, nor was a protocol prepared.

## SUPPLEMENT

**Supplemental figure S1.** OncoOrigin graphical user interface with initial window (a), inputted patient age, sex, and genes with detected variants (b), bottom of the window with predict function (c), and prediction report with predicted site, confidence, and relative probabilities for other sites (d).

**Supplemental table S2.** List of all genes included in data extraction from cBioPortal.

**Supplemental table S3.** List of all genes included in modeling after preprocessing and feature selection.

**Supplemental figure S4.** Class-specific precision-recall analysis curves for the RFC model (a), XGBC model (b), CBC model (c), and ETC model (d).

