## Supplementary figures and images for "Development and Validation of OncoOrigin: An Integrative AI Tool for Primary Cancer Site Prediction"

### Supplemental figure S1. OncoOrigin graphical user interface

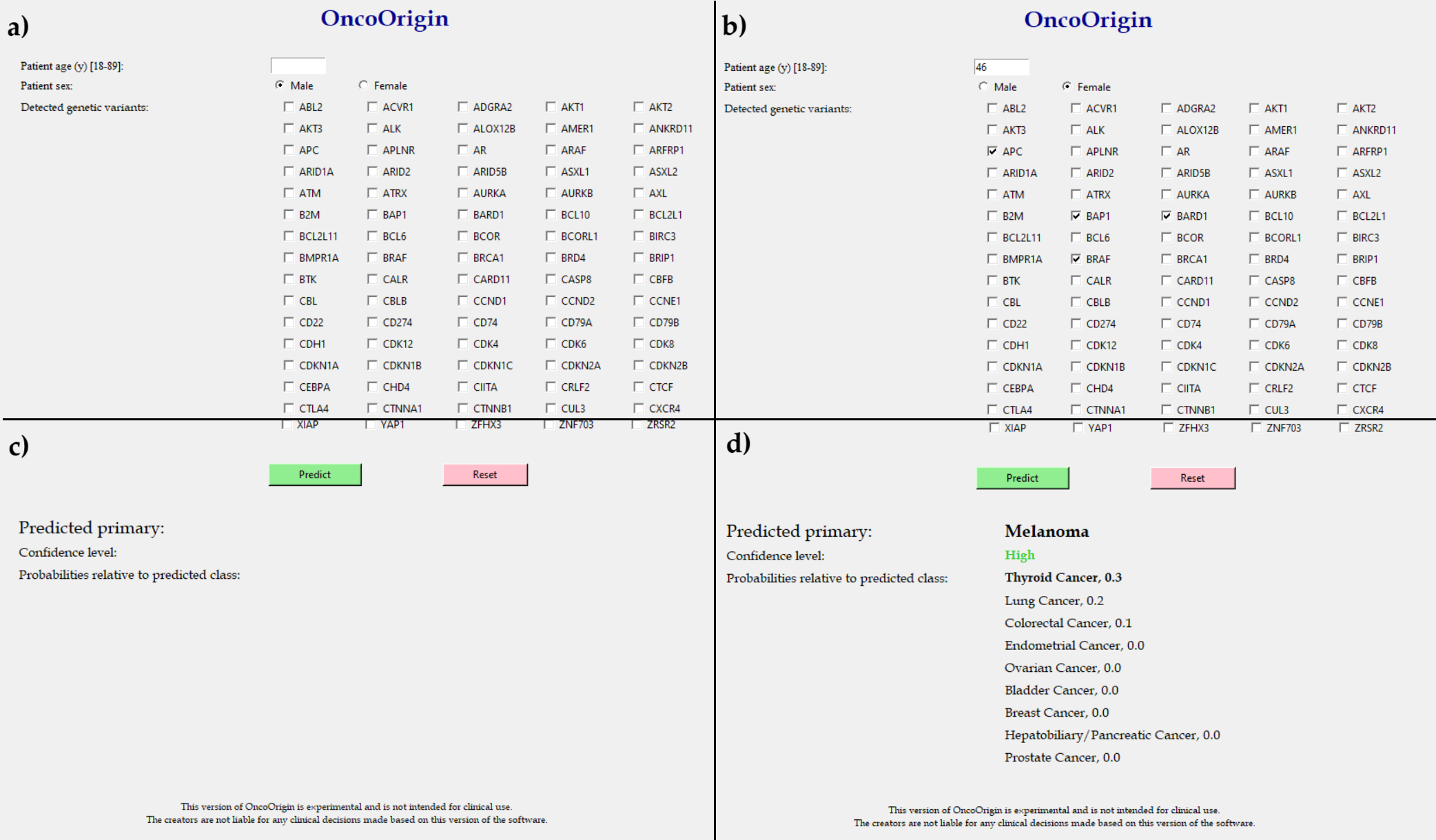

### Supplemental figure S4. Class-specific precision-recall analysis curves for the RFC model (a), XGBC model (b), CBC model (c), and ETC model (d)

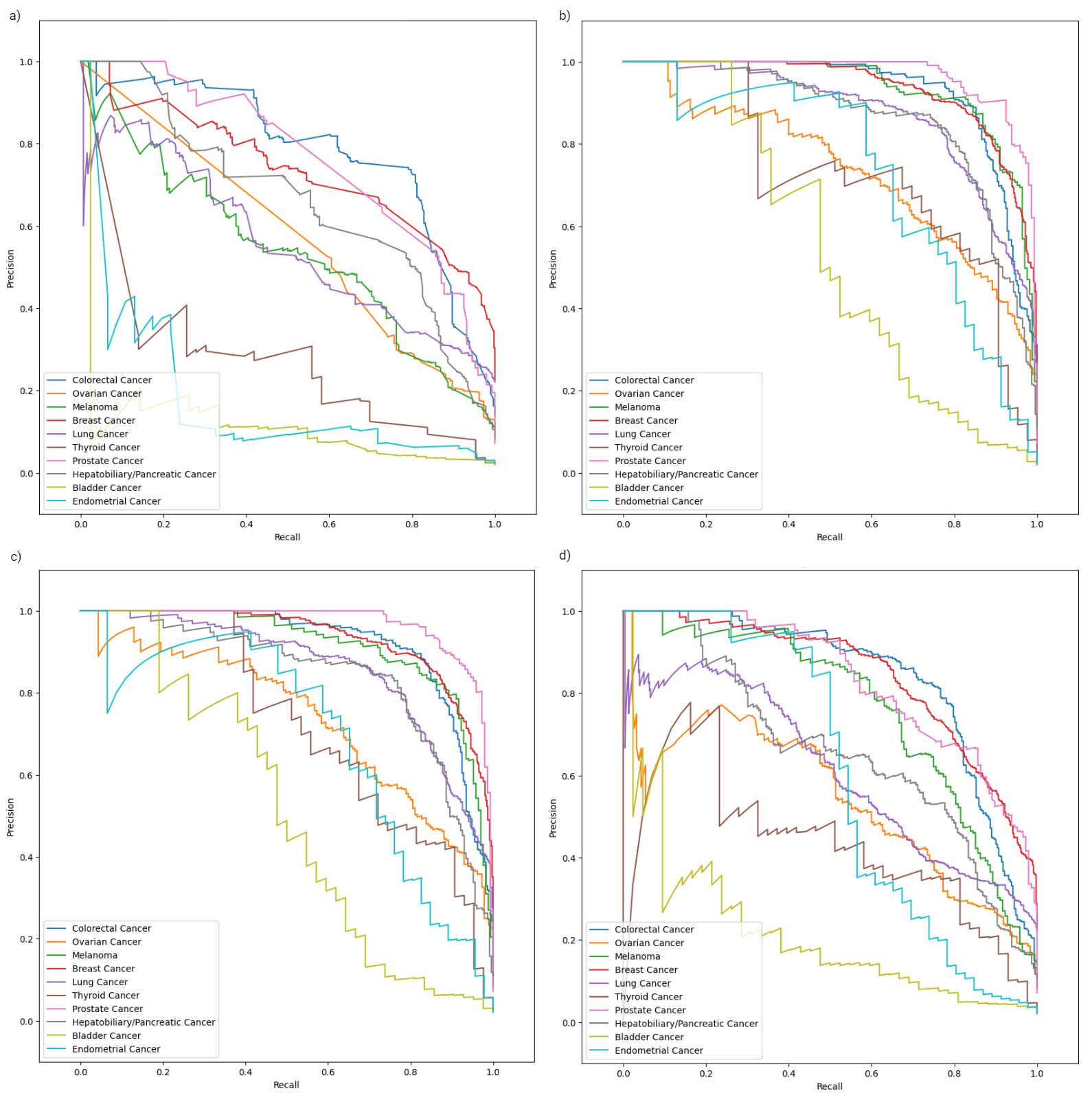
